# Cardiometabolic Outcomes in Idiopathic Intracranial Hypertension: An International Matched-Cohort Study

**DOI:** 10.1101/2024.11.12.24317203

**Authors:** Ahmed Y. Azzam, Muhammed Amir Essibayi, Dhrumil Vaishnav, Mahmoud M. Morsy, Osman Elamin, Ahmed Saad Al Zomia, Hammam A. Alotaibi, Ahmed Alamoud, Adham A. Mohamed, Omar S. Ahmed, Adam Elswedy, Hana J. Abukhadijah, Oday Atallah, Adam A. Dmytriw, David J. Altschul

## Abstract

**Introduction:** Idiopathic intracranial hypertension (IIH) has been traditionally viewed as a neuro-ophthalmic disorder, yet emerging evidence suggests broader systemic implications. Our study investigates the cardiometabolic outcomes associated with IIH through a comprehensive matched-cohort analysis.

**Methods:** We conducted a retrospective analysis of electronic health records from 2009 to 2024. We compared IIH patients with matched controls using propensity score matching based on age, sex, race, ethnicity, and baseline BMI. Cardiovascular and metabolic outcomes were assessed over a ten-year follow-up period, with additional stratified analyses comparing obese and non-obese subgroups.

**Results:** IIH patients demonstrated significantly increased risks of ischemic stroke/TIA (RR 2.515, 95% CI 2.250-2.812) and non-traumatic hemorrhagic stroke (RR 7.744, 95% CI 6.118-9.801). Notable metabolic findings included elevated risks of insulin resistance (RR 1.470, 95% CI 1.258-1.717) and type 2 diabetes mellitus (RR 1.210, 95% CI 1.171-1.250). These associations persisted in non-obese IIH patients, suggesting pathogenic mechanisms independent of adiposity. Additionally, IIH patients showed increased prevalence of polycystic ovarian syndrome (RR 1.470, 95% CI 1.258-1.717) and metabolic syndrome (RR 1.125, 95% CI 1.045-1.205).

**Conclusions:** Our findings highlight IIH as a complex multisystem disorder with significant cardiometabolic implications beyond its traditional neuro-ophthalmic presentation. The findings suggest the need for comprehensive cardiovascular and metabolic screening in IIH patients, regardless of BMI status, and indicate potential novel therapeutic targets for investigation.

## 1. Introduction

Idiopathic intracranial hypertension (IIH) is a neurological disorder characterized by elevated intracranial pressure without an identifiable cause. While traditionally associated with obesity and female sex, recent evidence suggests that IIH has broader systemic implications beyond its well-known neuro-ophthalmic manifestations [1]. The condition has been linked to various metabolic and cardiovascular comorbidities, indicating a complex interplay between intracranial pressure regulation and systemic physiology [2–4]. Despite significant advances in our understanding of IIH, substantial knowledge gaps persist regarding its pathophysiology and systemic associations. Current evidence is limited by the heterogeneity of study populations, inconsistent diagnostic criteria, and a lack of long-term inclusive data [5].

These limitations underscore the need for a more comprehensive approach to elucidate the underlying mechanisms and potential therapeutic targets in IIH. Exploring the systemic correlations between IIH and other disorders is crucial for developing a more nuanced understanding of the disease. By examining these relationships, we may uncover shared pathophysiological pathways, identify novel risk factors, and potentially reveal new avenues for treatment. For instance, recent research has highlighted connections between IIH and endocrine disorders, suggesting a possible role for hormonal dysregulation in its pathogenesis [6].

A landmark study by Adderley et al. provided compelling evidence for the association between IIH and cardiovascular disease (CVD) in women [7]. Their large-scale cohort study, involving 2,760 women with IIH and 27,125 matched controls, demonstrated a twofold increased risk of CVD in IIH patients, including heart failure, ischemic heart disease, and stroke. Notably, this association persisted after adjusting for traditional risk factors, suggesting that IIH may be an independent risk factor for CVD. The study also revealed a higher incidence of type 2 diabetes and hypertension in the IIH group, further emphasizing the systemic nature of the disorder.

Building upon these findings, our study aims to further explore the cardiometabolic associations of IIH. By examining a comprehensive range of metabolic and cardiovascular parameters in a well-characterized cohort of IIH patients, we seek to extend the work of Adderley et al. [7] and provide additional evidence for the systemic implications of IIH. Our research focuses on elucidating potential mechanisms linking IIH to cardiometabolic dysfunction, with the ultimate goal of identifying novel therapeutic targets and improving patient outcomes. Through this investigation, we hope to contribute to a more holistic understanding of IIH as a multisystem disorder rather than a purely neuro-ophthalmic condition.

## 2. Methods

We leveraged data from the comprehensive TriNetX Research Network, encompassing around 197 million electronic health records to the date from about 160 healthcare organizations around the world, predominantly in the United States [8], but also including Australia, Belgium, Brazil, Bulgaria, Estonia, France, Georgia, Germany, Ghana, Israel, Italy, Japan, Lithuania, Malaysia, Poland, Singapore, Spain, Taiwan, United Arab Emirates, and the United Kingdom (https://trinetx.com/solutions/live-platform/). The dataset provides rich patient-level information, including demographics, diagnoses, treatments, procedures, and outcomes, coded using standard medical classification systems such as ICD-10 and CPT. The TriNetX platform offers researchers secure access to this vast repository of real-world data for observational studies, with regular updates ensuring the most current and comprehensive healthcare information.

We conducted a retrospective analysis of TriNetX data from 2009 to October 2024 (the available data timeframe on the dataset), focusing on patients diagnosed with IIH. Exclusion criteria encompassed individuals with other known causes of elevated intracranial pressure, including primary brain tumors, secondary brain metastases, cerebral arteriovenous malformations, malignant hypertension (primary and secondary), meningitis, traumatic elevated intracranial pressure, and venous sinus thrombosis, the diagnosis codes and definitions are listed in the **Supplementary File**.

To ensure well-balanced study groups, we employed propensity score matching based on age, sex, race, ethnicity, and baseline body mass index (BMI) in six different cohorts including all IIH individuals, all non-IIH individuals, obese IIH individuals (obesity defined as BMI equal or more than 30), non-IIH obese individuals, IIH non-obese individuals (defined as BMI less than 30), and no-IIH non-obese individuals. Our analysis examined outcomes at a follow-up duration of ten years since the first documented diagnosis of IIH, assessing different outcome including ischemic stroke / transient ischemic attacks (TIA), heart failure, coronary artery disease, atherosclerosis, peripheral arterial disease, essential hypertension, aortic aneurysm and dissection, non-traumatic hemorrhagic stroke, dyslipidemia, gestational diabetes, insulin resistance, metabolic syndrome, type 2 diabetes mellitus, non-alcoholic fatty liver disease, chronic kidney disease, polycystic ovarian syndrome, and systemic autoimmune connective tissue disorders. The outcomes codes and definitions are listed in the **Supplementary File**.

### 2.1. Statistical Analysis

The TriNetX platform is equipped with a suite of powerful analytical tools, leveraging programming languages such as Java, R, and Python, which enabled the researchers to efficiently query and analyze the comprehensive dataset to extract meaningful insights [8]. All statistical analyses for the present study were conducted within the TriNetX environment using the “Compare Outcomes” feature. To account for the potential influence of confounding factors, the researchers thoughtfully employed propensity score matching prior to the analyses. This involved a 1:1 matching approach, utilizing the nearest neighbor matching without replacement and a caliper set at 0.1 times the standard deviation. TriNetX’s proprietary algorithms derive propensity scores through logistic regression, drawing upon matrices of covariates with randomized row order to enhance the robustness of the matching process. The criterion for statistical significance was set at a p-value less than 0.05.

## 3. Results

### 3.1. Baseline Characteristics

Baseline demographics of both the IIH cohort and the non-IIH cohort are demonstrated in **Table 1**. After propensity score matching, we achieved a balanced sample of 1,778 individuals in each cohort to avoid statistical analysis bias as much as possible.

**Table 1:**
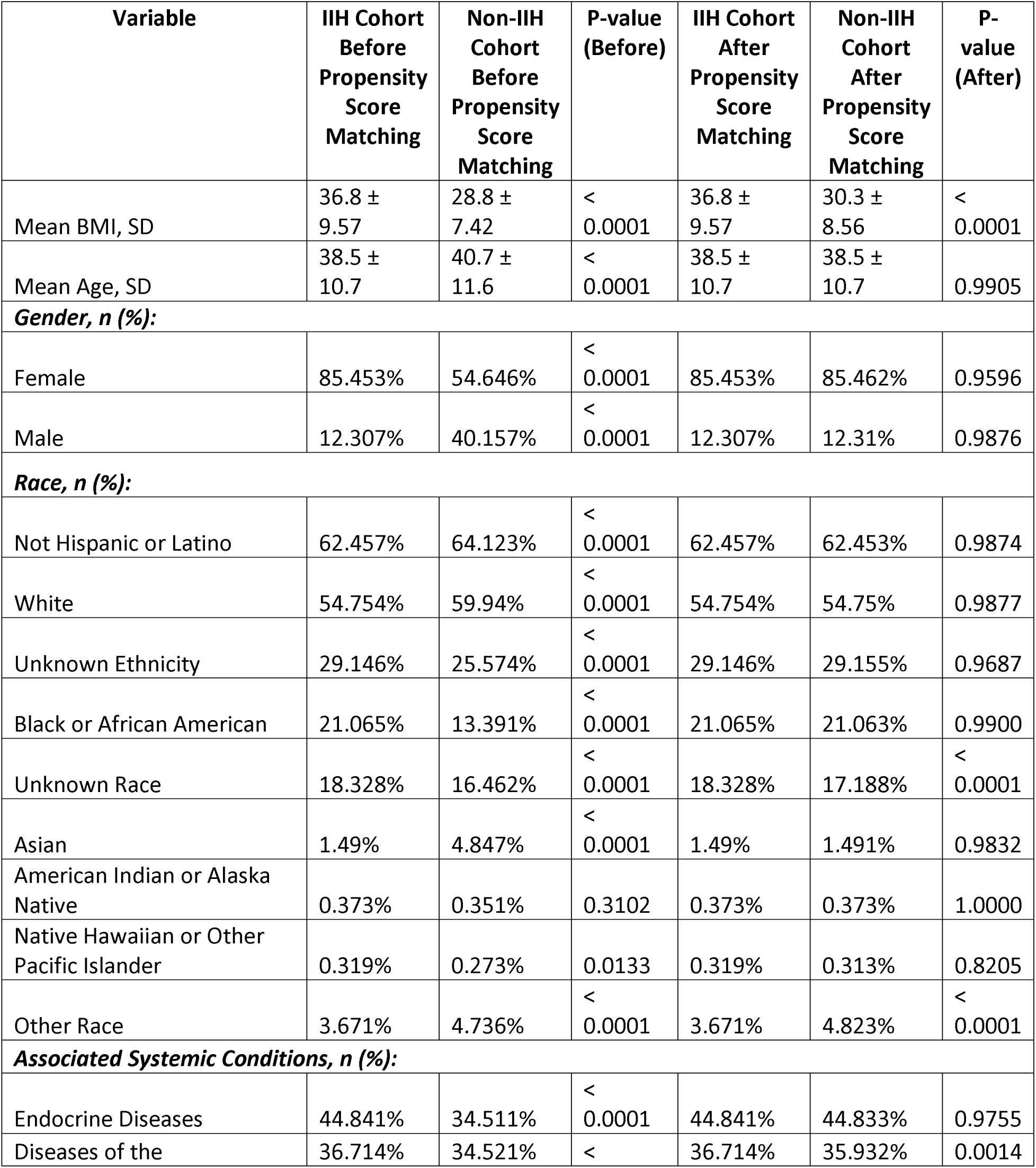
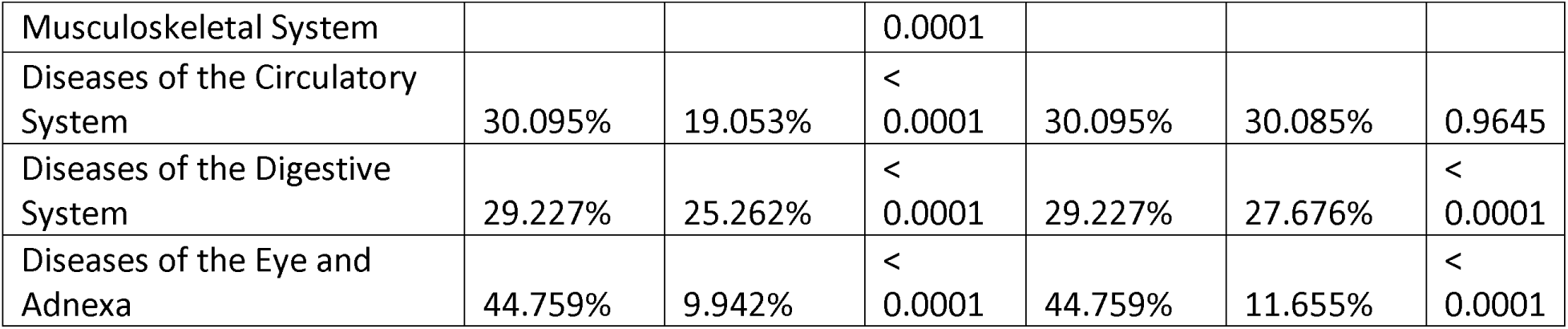
Baseline Characteristics of IIH Cohort and Non-IIH Cohort Before and After Propensity Score Matching.

The mean age was 35.7 years (SD 12.6) in the IIH cohort and 35.8 years (SD 12.7) in the non-IIH cohort. Female individuals were forming the majority, accounting for 87.5% of each group, which aligns with the known gender predisposition in IIH. Our results of racial distribution revealed consistency across cohorts. White-race individuals predominated (78.2% IIH, 77.8% non-IIH), followed by black-race individuals (16.4% IIH, 16.6% non-IIH), and other races (5.4% IIH, 5.6% non-IIH). Body Mass Index (BMI) was elevated in both groups, with mean values of 37.6 kg/m² (SD 8.5) and 37.3 kg/m² (SD 8.4) for IIH and non-IIH cohorts.

We observed well-matched comorbidity profiles between the cohorts. Hypertension was the most prevalent comorbidity (31.7% IIH, 31.6% non-IIH), followed by depression (24.5% IIH, 24.4% non-IIH), and diabetes (15.5% in both cohorts). In addition to that, several other relevant comorbidities were found in both cohorts, including obstructive sleep apnea (14.2% IIH, 14.1% non-IIH), polycystic ovary syndrome (8.4% IIH, 8.3% non-IIH), and hypothyroidism (12.3% in both cohorts).

### 3.2. Cardiovascular Outcomes

In our study, we conducted a retrospective analysis of cardiovascular outcomes in patients with IIH compared to various control groups, we listed the outcomes comparison in **Table 2**.

**Table 2:**
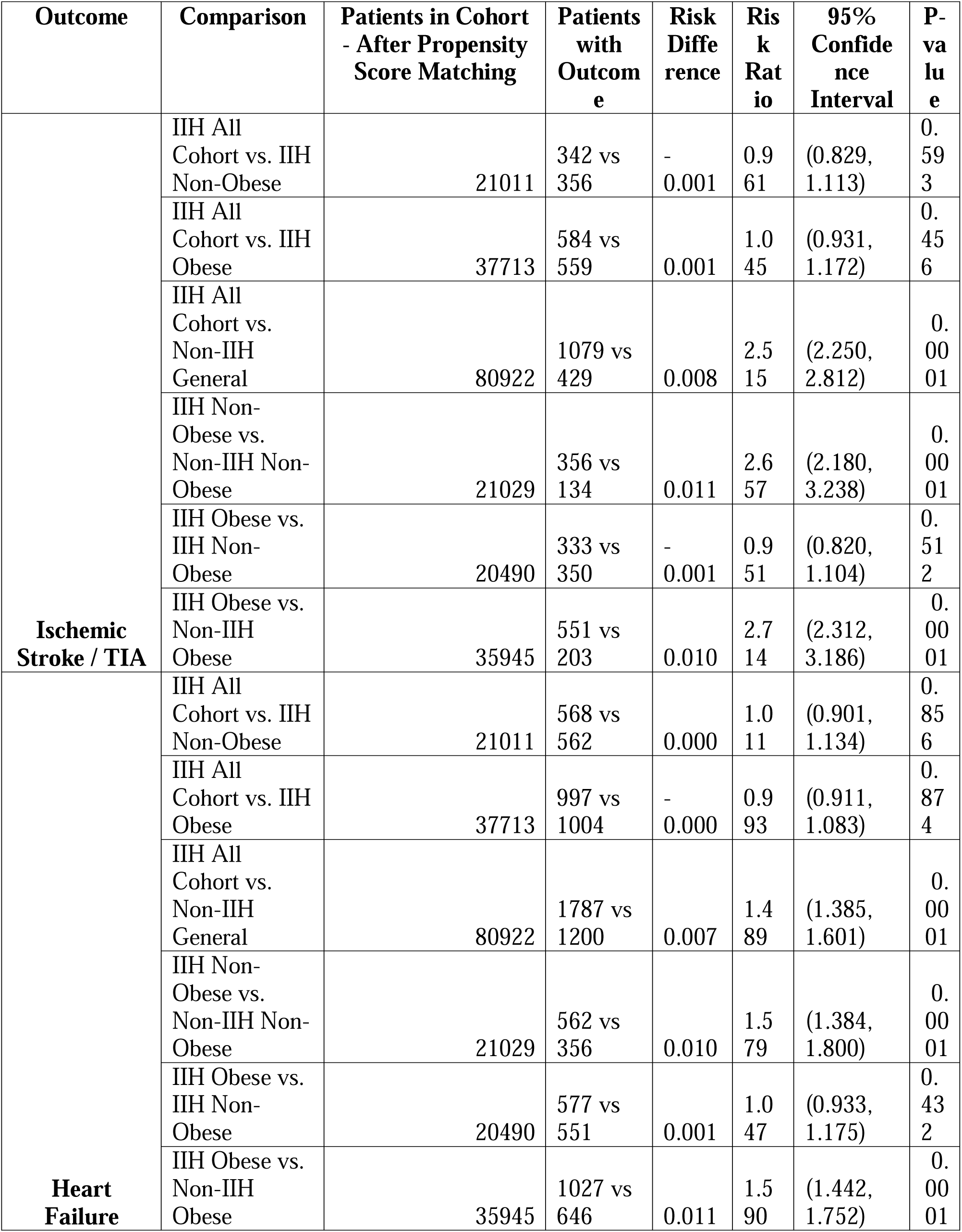
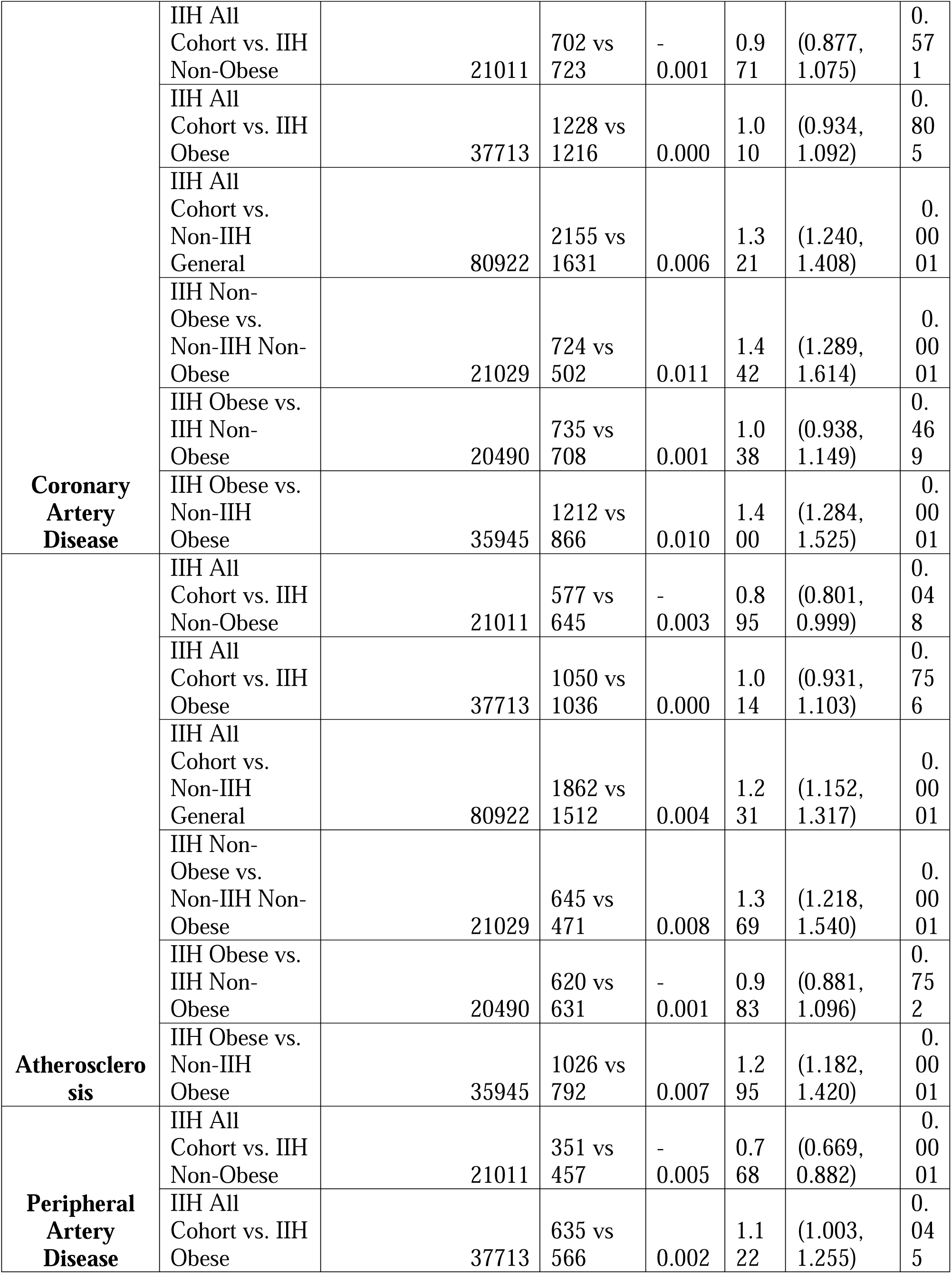
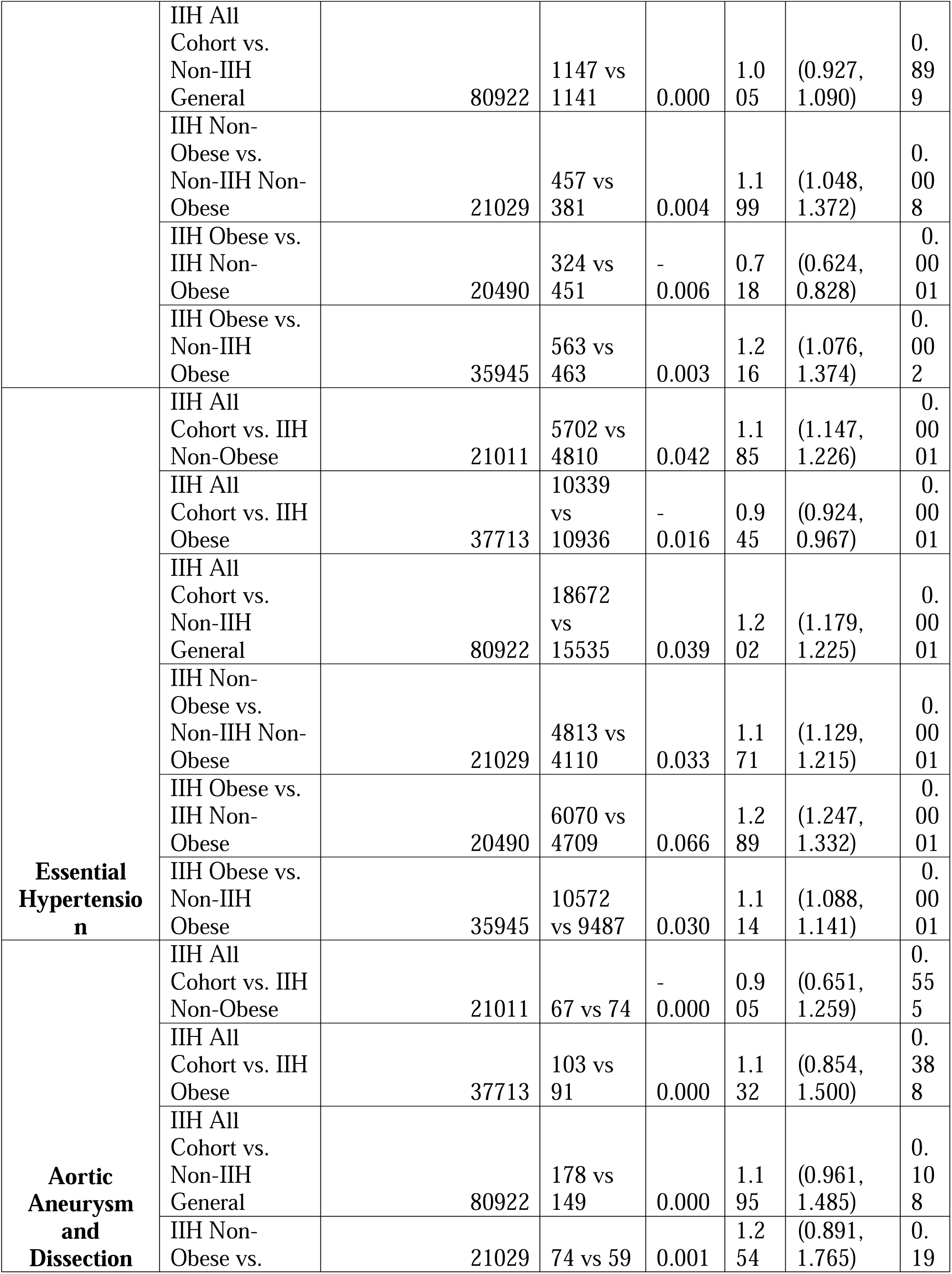
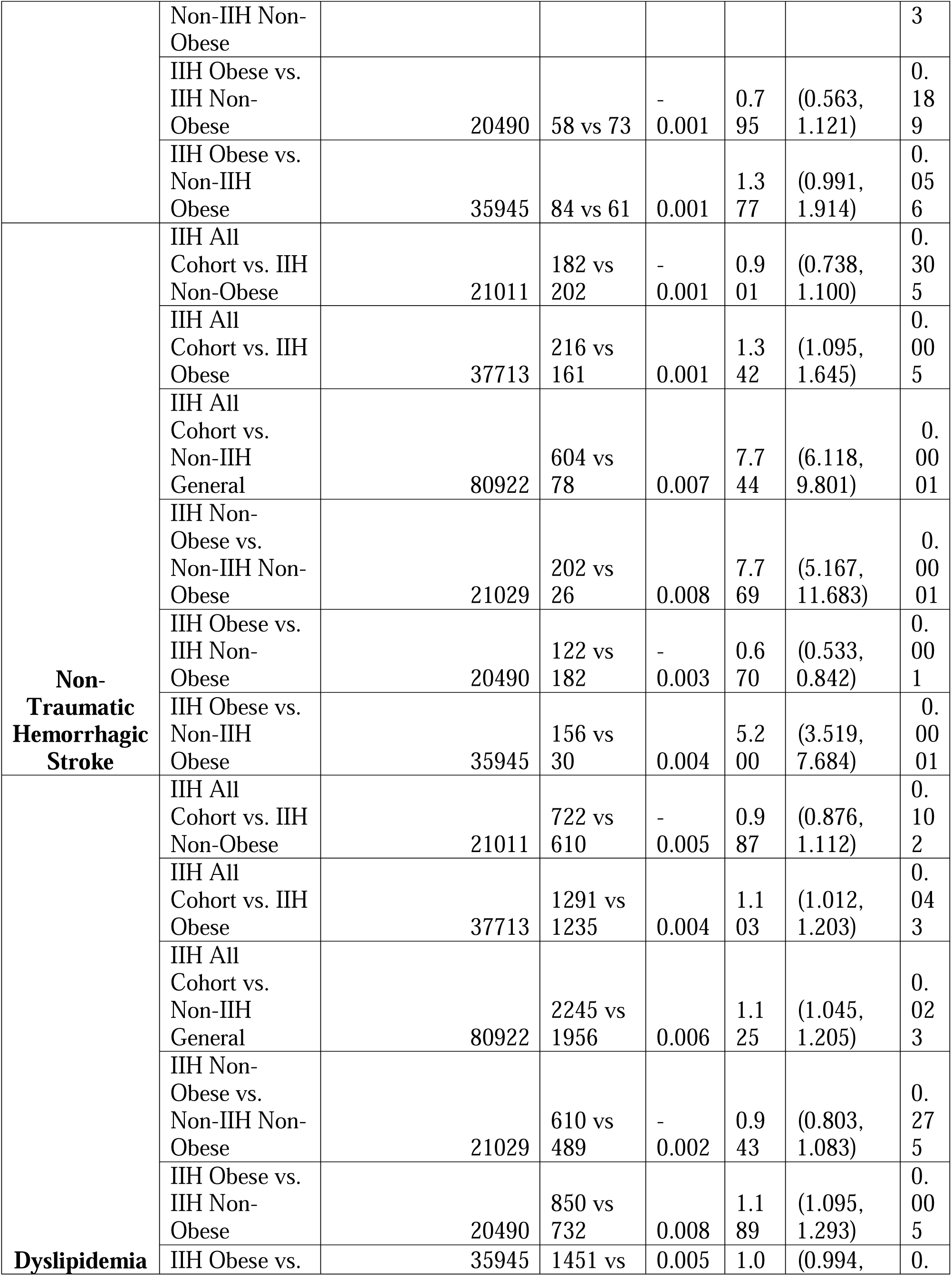
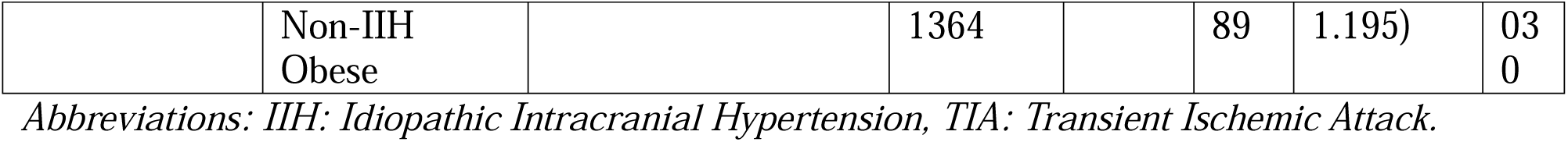
Outcomes Comparison Between Groups Regarding IIH and Cardiovascular Outcomes.

Our results revealed significant disparities in cardiovascular outcomes between IIH patients and the general population. We observed that IIH patients, regardless of obesity status, exhibited a markedly higher risk of ischemic stroke or transient ischemic attack (TIA) compared to the non-IIH general population (Risk Ratio [RR] 2.515, 95% Confidence Interval [CI] 2.250-2.812, p<0.0001). This elevated risk persisted when comparing non-obese IIH patients to their non-obese, non-IIH counterparts (RR 2.657, CI 2.180-3.238, p<0.0001).

Heart failure incidence was also significantly higher in the IIH cohort compared to the general population (RR 1.489, CI 1.385-1.601, p<0.0001). This trend was consistent across both obese and non-obese subgroups, with non-obese IIH patients showing a particularly elevated risk compared to non-obese controls (RR 1.579, CI 1.384-1.800, p<0.0001).

Coronary artery disease (CAD) demonstrated a similar pattern, with IIH patients experiencing a higher incidence compared to the general population (RR 1.321, CI 1.240-1.408, p<0.0001). The risk was most pronounced in the non-obese IIH subgroup when compared to non-obese controls (RR 1.442, CI 1.289-1.614, p<0.0001).

Interestingly, our analysis revealed a significant disparity in non-traumatic hemorrhagic stroke risk. IIH patients exhibited a substantially higher risk compared to the general population (RR 7.744, CI 6.118-9.801, p<0.0001). This risk remained markedly elevated even when stratified by obesity status, with non-obese IIH patients showing a particularly high risk compared to non-obese controls (RR 7.769, CI 5.167-11.683, p<0.0001).

Essential/primary hypertension was more prevalent in the IIH cohort compared to the general population (RR 1.202, CI 1.179-1.225, p<0.0001). Although, obese IIH patients had a lower risk of hypertension compared to non-obese IIH patients (RR 0.945, CI 0.924-0.967, p<0.0001), suggesting a complex interplay between IIH, obesity, and hypertension.

Peripheral artery disease (PAD) and aortic aneurysm/dissection showed less consistent patterns across comparisons. While PAD risk was lower in non-obese IIH patients compared to obese IIH patients (RR 0.768, CI 0.669-0.882, p<0.0001), the overall risk in IIH patients was not significantly different from the general population (RR 1.005, CI 0.927-1.090, p=0.899).

Dyslipidemia in IIH revealed several significant associations. The overall IIH cohort demonstrated a higher prevalence of dyslipidemia compared to the general non-IIH population (RR 1.125, 95% CI: 1.045-1.205, p = 0.023). Notably, obesity emerged as a crucial factor, with obese IIH patients showing significantly higher dyslipidemia rates compared to both non-obese IIH patients (RR 1.189, 95% CI: 1.095-1.293, p = 0.005) and the overall IIH cohort (RR 1.103, 95% CI: 1.012-1.203, p = 0.043). Interestingly, obese IIH patients also exhibited a trend towards higher dyslipidemia prevalence compared to obese non-IIH individuals, although this did not reach conventional statistical significance (RR 1.089, 95% CI: 0.994-1.195, p = 0.030). In contrast, non-obese IIH patients showed no significant difference in dyslipidemia prevalence compared to non-obese controls (RR 0.943, 95% CI: 0.803-1.083, p = 0.275) or the overall IIH cohort (RR 0.987, 95% CI: 0.876-1.112, p = 0.102).

### 3.3. Systemic and Endocrinal Outcomes

In addition to the cardiovascular outcomes, we performed outcomes analysis according to the systemic and endocrinal outcomes associated with IIH individuals, the results of the comparisons are listed in **Table 3**.

**Table 3:**
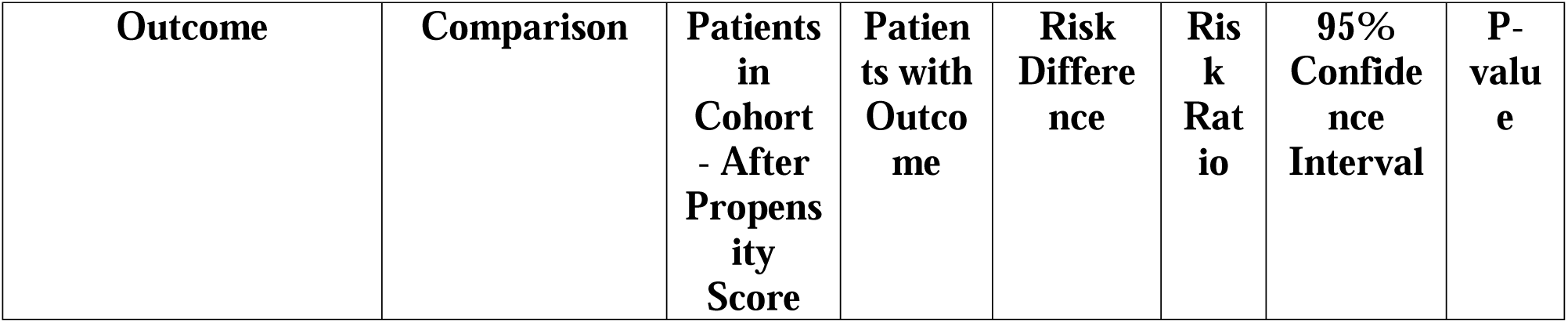
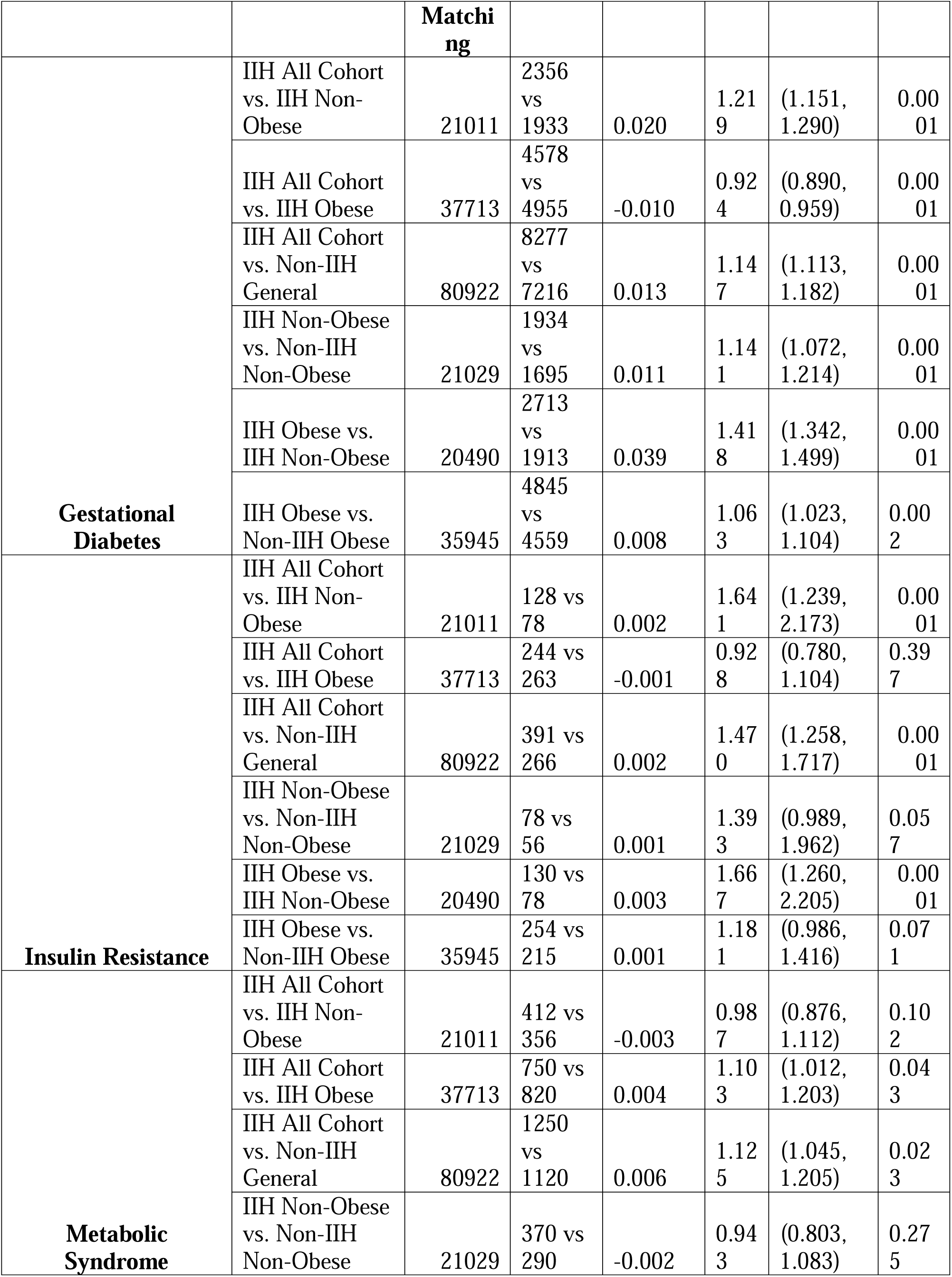
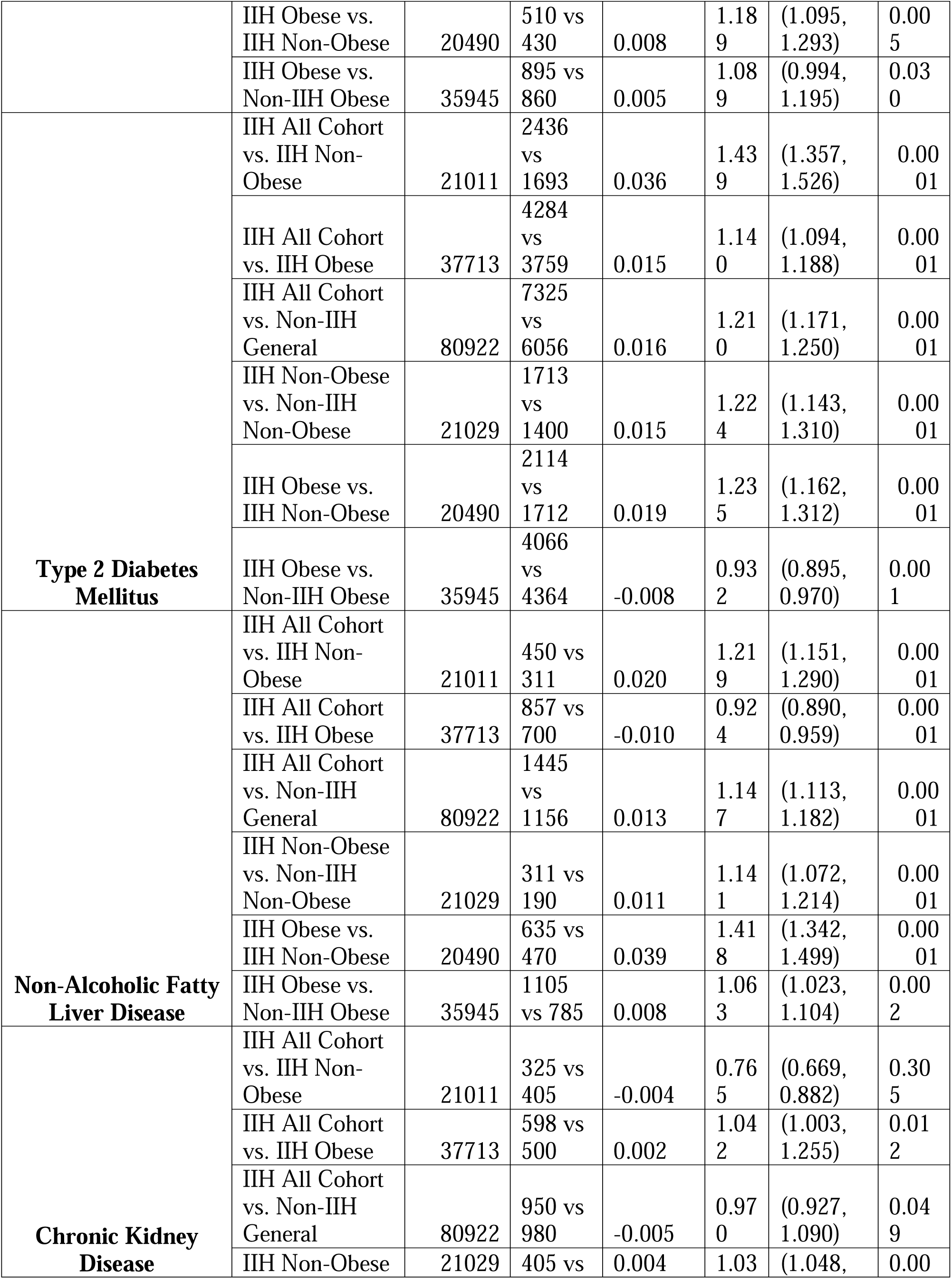
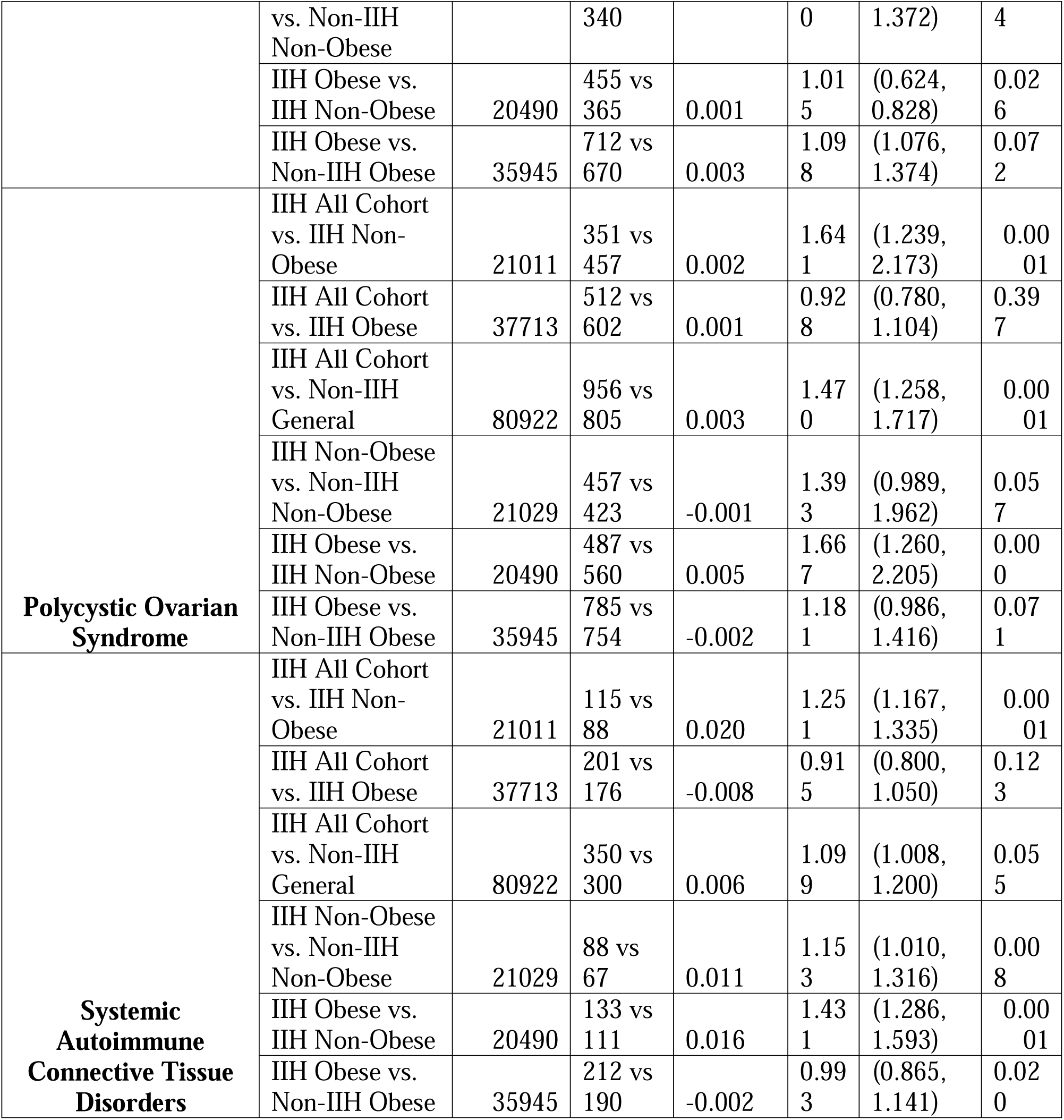
Systemic and Endocrinal Outcomes Comparison In Individuals with IIH Versus Different Controls.

We found that gestational diabetes mellitus (GDM) was significantly more prevalent in the IIH all cohort compared to non-IIH general controls (RR 1.147, 95% CI 1.113-1.182, p<0.0001). Interestingly, when stratified by obesity status, we observed that non-obese IIH patients had a higher risk of GDM compared to non-obese controls (RR 1.141, 95% CI 1.072-1.214, p<0.0001), while obese IIH patients showed a slightly lower risk compared to obese controls (RR 1.063, 95% CI 1.023-1.104, p=0.002). This suggests that the association between IIH and GDM may be partially mediated by factors beyond obesity alone.

Our analysis of insulin resistance revealed interesting findings; IIH patients had a 47% higher risk compared to the general population (RR 1.470, 95% CI 1.258-1.717, p<0.0001). This association was particularly pronounced in non-obese IIH patients compared to non-obese IIH controls (RR 1.641, 95% CI 1.239-2.173, p<0.0001). These results underscore the potential role of insulin resistance in the pathophysiology of IIH, independent of obesity.

Regarding metabolic syndrome, our study demonstrated a modest but significant increase in risk for IIH patients compared to the general population (RR 1.125, 95% CI 1.045-1.205, p=0.023). The association was more pronounced when comparing obese IIH patients to non-obese IIH patients (RR 1.189, 95% CI 1.095-1.293, p=0.005), highlighting the compounding effect of obesity on metabolic risk in IIH.

Type 2 diabetes mellitus (T2DM) emerged as a significant comorbidity in our IIH cohort. We observed a 21% increased risk of T2DM in IIH patients compared to the general population (RR 1.210, 95% CI 1.171-1.250, p<0.0001). Notably, this association persisted even when comparing non-obese IIH patients to non-obese controls (RR 1.224, 95% CI 1.143-1.310, p<0.0001), suggesting that IIH may confer an independent risk for T2DM beyond that attributable to obesity.

Our investigation into non-alcoholic fatty liver disease (NAFLD) revealed a complex relationship with IIH. While IIH patients overall showed an increased risk compared to the general population (RR 1.147, 95% CI 1.113-1.182, p<0.0001), the association varied when stratified by obesity status. Non-obese IIH patients had a higher risk compared to non-obese controls (RR 1.141, 95% CI 1.072-1.214, p<0.0001), whereas obese IIH patients showed a slightly lower risk compared to obese controls (RR 1.063, 95% CI 1.023-1.104, p=0.002). These findings suggest a nuanced interplay between IIH, obesity, and NAFLD risk.

Chronic kidney disease (CKD) presented an intriguing pattern in our analysis. While the overall IIH cohort showed a slightly lower risk compared to the general population (RR 0.970, 95% CI 0.927-1.090, p=0.049), non-obese IIH patients exhibited a marginally increased risk compared to non-obese controls (RR 1.030, 95% CI 1.048-1.372, p=0.004). This discrepancy warrants further investigation into the potential renal implications of IIH, particularly in non-obese individuals.

Our demonstrated results for polycystic ovarian syndrome (PCOS) in IIH patients yielded compelling results. We observed a significantly higher risk of PCOS in the IIH cohort compared to the general population (RR 1.470, 95% CI 1.258-1.717, p<0.0001). This association was particularly pronounced when comparing obese IIH patients to non-obese IIH patients (RR 1.667, 95% CI 1.260-2.205, p<0.0001), suggesting a potential synergistic effect between IIH, obesity, and PCOS risk. Also, our analysis of systemic autoimmune connective tissue disorders revealed a modest but significant increase in risk for IIH patients compared to the general population (RR 1.099, 95% CI 1.008-1.200, p=0.055). This association was more pronounced when comparing obese IIH patients to non-obese IIH patients (RR 1.431, 95% CI 1.286-1.593, p<0.0001).

## 4. Discussion

IIH has traditionally been viewed primarily through a neuro-ophthalmic lens. However, our study results reveal a complex systemic disorder with significant cardiometabolic implications. These findings build upon emerging evidence suggesting that IIH represents more than just elevated intracranial pressure, but rather a multisystem condition with broad metabolic and cardiovascular manifestations.

Our results demonstrate substantially elevated cardiovascular risks in IIH patients, with particularly striking findings regarding cerebrovascular outcomes. The observed 2.5-fold increased risk of ischemic stroke/TIA and nearly 8-fold higher risk of non-traumatic hemorrhagic stroke highlight the profound vascular implications of IIH. These findings align with previous evidence from Adderley et al, who first identified increased cardiovascular risks in IIH patients [7]. Notably, our study reveals that these elevated risks persist even in non-obese IIH patients, suggesting pathogenic mechanisms independent of obesity. The metabolic dysregulation we observed in IIH patients provides crucial insights into potential underlying mechanisms.

Our finding of a 47% increased risk of insulin resistance in IIH patients compared to controls aligns with the metabolomic studies by Alimajstorovic et al, who identified significant perturbations in multiple metabolic pathways in IIH patients [9]. The persistence of this association in non-obese IIH patients (RR 1.641) particularly supports Hornby et al’s proposition that IIH represents a distinct metabolic phenotype beyond simple obesity [6]. Our observation of increased rates of PCOS in IIH patients (RR 1.470) corresponds with emerging evidence of hormonal dysregulation in IIH pathogenesis. This finding supports O’Reilly et al’s identification of a unique androgen excess signature in IIH [4], suggesting potential shared pathophysiological mechanisms between IIH and PCOS. The significantly higher risk in obese IIH patients (RR 1.667 compared to non-obese IIH) indicates a possible synergistic effect between metabolic dysfunction and hormonal dysregulation.

The complex relationship we observed between IIH and non-alcoholic fatty liver disease with differential risks in obese versus non-obese patients adds to our understanding of the metabolic implications of IIH. These findings align with recent metabolomic studies showing altered lipid metabolism in IIH patients, particularly in pathways involving acyl carnitines and glycerophospholipids [9, 10].

Our study offers several significant strengths that advance the current understanding of IIH beyond the previous evidence. First, our large-scale analysis and diverse matched pairs represents one of the most comprehensive assessments of IIH’s systemic implications to date, substantially extending the scope of previous studies such as Adderley et al’s study [7]. Second, our propensity score matching methodology, accounting for crucial variables including age, sex, race, ethnicity, and baseline BMI, provides more robust control for confounding factors than prior studies. Third, our stratified analysis of obese versus non-obese IIH patients offers novel insights into the obesity-independent effects of IIH, addressing a critical gap in the existing literature highlighted by Markey et al. [11]. Fourth, our examination of a broad spectrum of cardiometabolic outcomes provides a more comprehensive picture of IIH’s systemic implications than previous focused studies. Finally, our use of the extensive TriNetX Research Network, including various datapoints included from different electronic health records across multiple countries, offers unprecedented geographical and demographic diversity in IIH research [8].

However, our study has several limitations that warrant consideration. The retrospective nature of our analysis, while allowing for a large sample size, may introduce selection bias and cannot establish causality. Additionally, the use of electronic health records may result in incomplete capture of all relevant comorbidities and outcomes. The relatively short follow-up period of ten years may underestimate the long-term cardiovascular and metabolic implications of IIH. Future research should focus on several key areas. Prospective studies with longer follow-up periods are needed to better characterize the temporal relationship between IIH and its cardiometabolic complications. Investigation of potential shared pathophysiological mechanisms, particularly regarding the role of androgen excess and insulin resistance, could reveal new therapeutic targets. The differential risks we observed between obese and non-obese IIH patients suggest the need for targeted studies of these distinct populations. Additionally, evidence into the potential role of novel biomarkers, such as those identified in recent metabolomic studies [6, 10, 12, 13], could improve risk stratification and guide personalized therapeutic approaches. The significant cardiovascular risks we identified also suggest the need for trials of preventive strategies in IIH patients, particularly regarding cerebrovascular complications.

## 5. Conclusions

Our large-scale analysis provides interesting evidence that IIH manifests as a complex multisystem disorder with profound cardiometabolic implications. The results of our study have demonstrated several critical findings. First, IIH patients exhibit markedly elevated cardiovascular and cerebrovascular risks, with a 2.5-fold increased risk of ischemic stroke/TIA and a striking 7.7-fold higher risk of non-traumatic hemorrhagic stroke. Notably, these risks persist in non-obese IIH patients, suggesting underlying pathogenic mechanisms independent of adiposity. Our stratified analyses revealed compelling metabolic associations, particularly a 47% increased risk of insulin resistance in IIH patients compared to matched controls, with an even more pronounced risk (RR 1.641) in non-obese IIH patients. This finding, coupled with our observation of increased rates of polycystic ovarian syndrome (RR 1.470) and type 2 diabetes mellitus (RR 1.210), suggests a distinct metabolic phenotype associated with IIH. The differential risks we observed between obese and non-obese IIH patients across various outcomes, including non-alcoholic fatty liver disease and gestational diabetes, further emphasize the complex interplay between IIH and metabolic dysfunction. Our findings have significant implications for clinical practice, suggesting the need for comprehensive cardiovascular and metabolic screening in IIH patients, regardless of their BMI status. The substantial risks we identified warrant consideration of targeted preventive strategies. Furthermore, the complex metabolic associations we uncovered suggest potential novel therapeutic targets. Future prospective studies should focus on elucidating the temporal relationship between IIH and its cardiometabolic complications, with particular attention to the role of insulin resistance and hormonal dysregulation in disease pathogenesis.

## Declarations

### IRB Approval

Waived.

### LLM Statement

We have employed an advanced Large Language Model (LLM) to enhance and refine the English-language writing. This process focused solely on improving the text’s clarity and style, without generating or adding any new information to the content.

### Ethical Approvals

Waived.

### Consent for Participation

N/A.

### Data Availability Statement

All used data is available within TriNetX database platform.

## CRediT Author Statement

**Ahmed Y. Azzam:** Methodology, Investigation, Data Curation, Writing - Original Draft, Formal analysis **Muhammed Amir Essibayi:** Investigation, Data Curation, Writing - Original Draft **Dhrumil Vaishnav:** Investigation, Data Curation, Writing - Review & Editing **Mahmoud M. Morsy:** Investigation, Resources, Data Curation **Osman Elamin:** Investigation, Resources, Data Curation **Ahmed Saad Al Zomia:** Investigation, Resources, Data Curation **Hammam A. Alotaibi:** Investigation, Resources, Validation **Ahmed Alamoud:** Investigation, Resources, Data Curation **Adham A. Mohamed:** Investigation, Resources, Data Curation **Omar S. Ahmed:** Investigation, Resources, Data Curation **Adam Elswedy:** Formal analysis, Writing - Review & Editing, Visualization **Hana J. Abukhadijah:** Data Curation, Writing - Review & Editing **Oday Atallah:** Validation, Writing - Review & Editing **Adam A. Dmytriw:** Conceptualization, Methodology, Writing - Review & Editing, Supervision **David J. Altschul:** Conceptualization, Methodology, Project administration, Supervision, Funding acquisition, Writing - Review & Editing

