## Supplementary File for "Cardiometabolic Outcomes in Idiopathic Intracranial Hypertension: An International Matched-Cohort Study"

##### **Query Criteria for idiopathic intracranial hypertension (IIH):**

This query was run on the network Global Collaborative Network with 130 HCO(s) queried and 130 HCO(s) responded. A total of 125 provider(s) responded with patients. The final cohort included 83,384 patients who matched the query criteria listed in the table below. For the text representation of the query criteria please see Appendix A.

| Ungrouped terms | | | | | |
| --- | --- | --- | --- | --- | --- |
|  | must have |  | demographics | Age | Age (between 18 and 60 years (most recent occurrence)) |
|  |  | and | diagnosis | UMLS:ICD10CM:G93.2 | Benign intracranial hypertension |
|  | cannot have |  | diagnosis | UMLS:ICD10CM:Q28.2 | Arteriovenous malformation of cerebral vessels |
|  |  | or | diagnosis | UMLS:ICD10CM:G93.5 | Compression of brain |
|  |  | or | diagnosis | UMLS:ICD10CM:Q00-Q07 | Congenital malformations of the nervous system |
|  |  | or | diagnosis | UMLS:ICD10CM:G91 | Hydrocephalus |
|  |  | or | diagnosis | UMLS:ICD10CM:G03 | Meningitis due to other and unspecified causes |
|  |  | or | diagnosis | UMLS:ICD10CM:G08 | Intracranial and intraspinal phlebitis and thrombophlebitis |
|  |  | or | globaloncology | UMLS:ICDO3:C71 | Brain |
|  |  | or | diagnosis | UMLS:ICD10CM:C79.31 | Secondary malignant neoplasm of brain |

##### **Outcome Definitions:**

The table below outlines the definitions for each outcome and the analysis specifications. For outcome definitions consisting of more than one term, at least one term must match. Please see Appendix B for the text representation of the outcome definitions.

| Ischemic Stroke / TIA | | | | |
| --- | --- | --- | --- | --- |
|  | **Outcome definition** | | | |
|  | | Diagnosis | UMLS:ICD10CM:I63.50 | Cerebral infarction due to unspecified occlusion or stenosis of unspecified cerebral artery |
|  | | Diagnosis | UMLS:ICD10CM:G45 | Transient cerebral ischemic attacks and related syndromes |
|  | | Diagnosis | UMLS:ICD10CM:I67.848 | Other cerebrovascular vasospasm and vasoconstriction |
|  | | Diagnosis | UMLS:ICD10CM:I63.50 | Cerebral infarction due to unspecified occlusion or stenosis of unspecified cerebral artery |
|  | **Settings for the performed analyses** | | | |
|  | | Risk analysis | | including patients with outcome prior to the time window |
| Heart Failure | | | | |
|  | **Outcome definition** | | | |
|  | | Diagnosis | UMLS:ICD10CM:I50 | Heart failure |
|  | **Settings for the performed analyses** | | | |
|  | | Risk analysis | | including patients with outcome prior to the time window |
| Coronary Artery Disease | | | | |
|  | **Outcome definition** | | | |
|  | | Diagnosis | UMLS:ICD10CM:I20.0 | Unstable angina |
|  | | Diagnosis | UMLS:ICD10CM:I21.3 | ST elevation (STEMI) myocardial infarction of unspecified site |
|  | | Diagnosis | UMLS:ICD10CM:I21.29 | ST elevation (STEMI) myocardial infarction involving other sites |
|  | | Diagnosis | UMLS:ICD10CM:I21 | Acute myocardial infarction |
|  | | Diagnosis | UMLS:ICD10CM:I21.4 | Non-ST elevation (NSTEMI) myocardial infarction |
|  | | Diagnosis | UMLS:ICD10CM:I20 | Angina pectoris |
|  | | Diagnosis | UMLS:ICD10CM:I20.8 | Other forms of angina pectoris |
|  | | Diagnosis | UMLS:ICD10CM:I25.1 | Atherosclerotic heart disease of native coronary artery |
|  | | Diagnosis | UMLS:ICD10CM:I25.10 | Atherosclerotic heart disease of native coronary artery without angina pectoris |
|  | **Settings for the performed analyses** | | | |
|  | | Risk analysis | | including patients with outcome prior to the time window |
| Atherosclerosis | | | | |
|  | **Outcome definition** | | | |
|  | | Diagnosis | UMLS:ICD10CM:I70 | Atherosclerosis |
|  | | Diagnosis | UMLS:ICD10CM:I70.0 | Atherosclerosis of aorta |
|  | | Diagnosis | UMLS:ICD10CM:I25.1 | Atherosclerotic heart disease of native coronary artery |
|  | | Diagnosis | UMLS:ICD10CM:I25.10 | Atherosclerotic heart disease of native coronary artery without angina pectoris |
|  | | Diagnosis | UMLS:ICD10CM:I25.11 | Atherosclerotic heart disease of native coronary artery with angina pectoris |
|  | | Diagnosis | UMLS:ICD10CM:I70.2 | Atherosclerosis of native arteries of the extremities |
|  | | Diagnosis | UMLS:ICD10CM:I70.21 | Atherosclerosis of native arteries of extremities with intermittent claudication |
|  | | Diagnosis | UMLS:ICD10CM:I70.8 | Atherosclerosis of other arteries |
|  | | Diagnosis | UMLS:ICD10CM:I25.810 | Atherosclerosis of coronary artery bypass graft(s) without angina pectoris |
|  | | Diagnosis | UMLS:ICD10CM:I25.81 | Atherosclerosis of other coronary vessels without angina pectoris |
|  | | Diagnosis | UMLS:ICD10CM:I25.110 | Atherosclerotic heart disease of native coronary artery with unstable angina pectoris |
|  | **Settings for the performed analyses** | | | |
|  | | Risk analysis | | including patients with outcome prior to the time window |
| Essential Hypertension | | | | |
|  | **Outcome definition** | | | |
|  | | Diagnosis | UMLS:ICD10CM:I10 | Essential (primary) hypertension |
|  | **Settings for the performed analyses** | | | |
|  | | Risk analysis | | including patients with outcome prior to the time window |
| Type 2 Diabetes Mellitus | | | | |
|  | **Outcome definition** | | | |
|  | | Diagnosis | UMLS:ICD10CM:E11 | Type 2 diabetes mellitus |
|  | **Settings for the performed analyses** | | | |
|  | | Risk analysis | | including patients with outcome prior to the time window |
| Aortic Aneurysm and Dissection | | | | |
|  | **Outcome definition** | | | |
|  | | Diagnosis | UMLS:ICD10CM:I71 | Aortic aneurysm and dissection |
|  | **Settings for the performed analyses** | | | |
|  | | Risk analysis | | including patients with outcome prior to the time window |
| Non-Traumatic Hemorrhagic Stroke | | | | |
|  | **Outcome definition** | | | |
|  | | Diagnosis | UMLS:ICD10CM:I61 | Nontraumatic intracerebral hemorrhage |
|  | **Settings for the performed analyses** | | | |
|  | | Risk analysis | | including patients with outcome prior to the time window |
| Peripheral Artery Disease | | | | |
|  | **Outcome definition** | | | |
|  | | Diagnosis | UMLS:ICD10CM:I73 | Other peripheral vascular diseases |
|  | | Diagnosis | UMLS:ICD10CM:I73 | Other peripheral vascular diseases |
|  | | Diagnosis | UMLS:ICD10CM:I70.213 | Atherosclerosis of native arteries of extremities with intermittent claudication, bilateral legs |
|  | | Diagnosis | UMLS:ICD10CM:I70.218 | Atherosclerosis of native arteries of extremities with intermittent claudication, other extremity |
|  | | Diagnosis | UMLS:ICD10CM:I70.212 | Atherosclerosis of native arteries of extremities with intermittent claudication, left leg |
|  | | Diagnosis | UMLS:ICD10CM:I70.211 | Atherosclerosis of native arteries of extremities with intermittent claudication, right leg |
|  | | Diagnosis | UMLS:ICD10CM:I70.219 | Atherosclerosis of native arteries of extremities with intermittent claudication, unspecified extremity |
|  | | Diagnosis | UMLS:ICD10CM:I70.21 | Atherosclerosis of native arteries of extremities with intermittent claudication |
|  | **Settings for the performed analyses** | | | |
|  | | Risk analysis | | including patients with outcome prior to the time window |
| Gestational Diabetes | | | | |
|  | **Outcome definition** | | | |
|  | | Diagnosis | UMLS:ICD10CM:O24.4 | Gestational diabetes mellitus |
|  | | Diagnosis | UMLS:ICD10CM:E11 | Type 2 diabetes mellitus |
|  | | Diagnosis | UMLS:ICD10CM:O99.810 | Abnormal glucose complicating pregnancy |
|  | | Diagnosis | UMLS:ICD10CM:O24.911 | Unspecified diabetes mellitus in pregnancy, first trimester |
|  | | Diagnosis | UMLS:ICD10CM:O24.919 | Unspecified diabetes mellitus in pregnancy, unspecified trimester |
|  | | Diagnosis | UMLS:ICD10CM:O24.419 | Gestational diabetes mellitus in pregnancy, unspecified control |
|  | | Diagnosis | UMLS:ICD10CM:O24.429 | Gestational diabetes mellitus in childbirth, unspecified control |
|  | | Diagnosis | UMLS:ICD10CM:O24.439 | Gestational diabetes mellitus in the puerperium, unspecified control |
|  | | Diagnosis | UMLS:ICD10CM:O24.41 | Gestational diabetes mellitus in pregnancy |
|  | | Diagnosis | UMLS:ICD10CM:O24.42 | Gestational diabetes mellitus in childbirth |
|  | | Diagnosis | UMLS:ICD10CM:O24.43 | Gestational diabetes mellitus in the puerperium |
|  | | Diagnosis | UMLS:ICD10CM:O24.410 | Gestational diabetes mellitus in pregnancy, diet controlled |
|  | | Diagnosis | UMLS:ICD10CM:O24.414 | Gestational diabetes mellitus in pregnancy, insulin controlled |
|  | | Diagnosis | UMLS:ICD10CM:O24.420 | Gestational diabetes mellitus in childbirth, diet controlled |
|  | | Diagnosis | UMLS:ICD10CM:O24.424 | Gestational diabetes mellitus in childbirth, insulin controlled |
|  | | Diagnosis | UMLS:ICD10CM:O24.430 | Gestational diabetes mellitus in the puerperium, diet controlled |
|  | | Diagnosis | UMLS:ICD10CM:O24.434 | Gestational diabetes mellitus in the puerperium, insulin controlled |
|  | | Diagnosis | UMLS:ICD10CM:O99.81 | Abnormal glucose complicating pregnancy, childbirth and the puerperium |
|  | **Settings for the performed analyses** | | | |
|  | | Risk analysis | | including patients with outcome prior to the time window |
| Insulin Resistance | | | | |
|  | **Outcome definition** | | | |
|  | | Diagnosis | UMLS:ICD10CM:E88.819 | Insulin resistance, unspecified |
|  | | Diagnosis | UMLS:ICD10CM:E88.818 | Other insulin resistance |
|  | | Diagnosis | UMLS:ICD10CM:E88.811 | Insulin resistance syndrome, Type A |
|  | | Diagnosis | UMLS:ICD10CM:E72.52 | Trimethylaminuria |
|  | **Settings for the performed analyses** | | | |
|  | | Risk analysis | | including patients with outcome prior to the time window |
| Metabolic Syndrome | | | | |
|  | **Outcome definition** | | | |
|  | | Diagnosis | UMLS:ICD10CM:E88.810 | Metabolic syndrome |
|  | | Diagnosis | UMLS:ICD10CM:E88.81 | Metabolic syndrome and other insulin resistance |
|  | **Settings for the performed analyses** | | | |
|  | | Risk analysis | | including patients with outcome prior to the time window |
| Dyslipidemia | | | | |
|  | **Outcome definition** | | | |
|  | | Diagnosis | UMLS:ICD10CM:E78.5 | Hyperlipidemia, unspecified |
|  | | Diagnosis | UMLS:ICD10CM:E78.4 | Other hyperlipidemia |
|  | | Diagnosis | UMLS:ICD10CM:E78.5 | Hyperlipidemia, unspecified |
|  | | Diagnosis | UMLS:ICD10CM:E78.4 | Other hyperlipidemia |
|  | | Diagnosis | UMLS:ICD10CM:E78.0 | Pure hypercholesterolemia |
|  | | Diagnosis | UMLS:ICD10CM:E78.2 | Mixed hyperlipidemia |
|  | | Diagnosis | UMLS:ICD10CM:E78.1 | Pure hyperglyceridemia |
|  | | Diagnosis | UMLS:ICD10CM:E78.3 | Hyperchylomicronemia |
|  | | Diagnosis | UMLS:ICD10CM:E78.49 | Other hyperlipidemia |
|  | | Diagnosis | UMLS:ICD10CM:E78 | Disorders of lipoprotein metabolism and other lipidemias |
|  | **Settings for the performed analyses** | | | |
|  | | Risk analysis | | including patients with outcome prior to the time window |
| NAFLD | | | | |
|  | **Outcome definition** | | | |
|  | | Diagnosis | UMLS:ICD10CM:K76.0 | Fatty (change of) liver, not elsewhere classified |
|  | | Diagnosis | UMLS:ICD10CM:K75.81 | Nonalcoholic steatohepatitis (NASH) |
|  | | Diagnosis | UMLS:ICD10CM:K74 | Fibrosis and cirrhosis of liver |
|  | | Diagnosis | UMLS:ICD10CM:K74.0 | Hepatic fibrosis |
|  | **Settings for the performed analyses** | | | |
|  | | Risk analysis | | including patients with outcome prior to the time window |
| CKD | | | | |
|  | **Outcome definition** | | | |
|  | | Diagnosis | UMLS:ICD10CM:N18 | Chronic kidney disease (CKD) |
|  | | Diagnosis | UMLS:ICD10CM:N18.6 | End stage renal disease |
|  | | Diagnosis | UMLS:ICD10CM:N18.9 | Chronic kidney disease, unspecified |
|  | | Diagnosis | UMLS:ICD10CM:N18.3 | Chronic kidney disease, stage 3 (moderate) |
|  | | Diagnosis | UMLS:ICD10CM:N18.4 | Chronic kidney disease, stage 4 (severe) |
|  | | Diagnosis | UMLS:ICD10CM:N18.5 | Chronic kidney disease, stage 5 |
|  | | Diagnosis | UMLS:ICD10CM:N18.1 | Chronic kidney disease, stage 1 |
|  | **Settings for the performed analyses** | | | |
|  | | Risk analysis | | including patients with outcome prior to the time window |
| PCOS | | | | |
|  | **Outcome definition** | | | |
|  | | Diagnosis | UMLS:ICD10CM:E28.2 | Polycystic ovarian syndrome |
|  | **Settings for the performed analyses** | | | |
|  | | Risk analysis | | including patients with outcome prior to the time window |
| Systemic Autoimmune Connective Tissue Disorders | | | | |
|  | **Outcome definition** | | | |
|  | | Diagnosis | UMLS:ICD10CM:M30-M36 | Systemic connective tissue disorders |
|  | **Settings for the performed analyses** | | | |
|  | | Risk analysis | | including patients with outcome prior to the time window |

### **Appendix A – Text Representation of the Cohorts Definition:**

This section lists all terms used in the definitions of the two cohorts.

##### Query Criteria for IIH:

Patients must have:
 all of the following:
 Age (Age) (between 18 and 60 years (most recent occurrence)); and
 Benign intracranial hypertension (UMLS:ICD10CM:G93.2).

Patients cannot have:
 any of the following:
 Arteriovenous malformation of cerebral vessels (UMLS:ICD10CM:Q28.2); or
 Compression of brain (UMLS:ICD10CM:G93.5); or
 Congenital malformations of the nervous system (UMLS:ICD10CM:Q00-Q07); or
 Hydrocephalus (UMLS:ICD10CM:G91); or
 Meningitis due to other and unspecified causes (UMLS:ICD10CM:G03); or
 Intracranial and intraspinal phlebitis and thrombophlebitis (UMLS:ICD10CM:G08); or
 Brain (UMLS:ICDO3:C71); or
 Secondary malignant neoplasm of brain (UMLS:ICD10CM:C79.31).

### **Appendix B – Text Representation of the Outcomes Definition:**

This analysis includes the following outcomes:

Ischemic Stroke / TIA
 Patients must have:
 any of the following:
 Cerebral infarction due to unspecified occlusion or stenosis of unspecified cerebral artery (UMLS:ICD10CM:I63.50); or
 Transient cerebral ischemic attacks and related syndromes (UMLS:ICD10CM:G45); or
 Other cerebrovascular vasospasm and vasoconstriction (UMLS:ICD10CM:I67.848); or
 Cerebral infarction due to unspecified occlusion or stenosis of unspecified cerebral artery (UMLS:ICD10CM:I63.50).

Heart Failure
 Patients must have:
 Heart failure (UMLS:ICD10CM:I50).

Coronary Artery Disease
 Patients must have:
 any of the following:
 Unstable angina (UMLS:ICD10CM:I20.0); or
 ST elevation (STEMI) myocardial infarction of unspecified site (UMLS:ICD10CM:I21.3); or
 ST elevation (STEMI) myocardial infarction involving other sites (UMLS:ICD10CM:I21.29); or
 Acute myocardial infarction (UMLS:ICD10CM:I21); or
 Non-ST elevation (NSTEMI) myocardial infarction (UMLS:ICD10CM:I21.4); or
 Angina pectoris (UMLS:ICD10CM:I20); or
 Other forms of angina pectoris (UMLS:ICD10CM:I20.8); or
 Atherosclerotic heart disease of native coronary artery (UMLS:ICD10CM:I25.1); or
 Atherosclerotic heart disease of native coronary artery without angina pectoris (UMLS:ICD10CM:I25.10).

Atherosclerosis
 Patients must have:
 any of the following:
 Atherosclerosis (UMLS:ICD10CM:I70); or
 Atherosclerosis of aorta (UMLS:ICD10CM:I70.0); or
 Atherosclerotic heart disease of native coronary artery (UMLS:ICD10CM:I25.1); or
 Atherosclerotic heart disease of native coronary artery without angina pectoris (UMLS:ICD10CM:I25.10); or
 Atherosclerotic heart disease of native coronary artery with angina pectoris (UMLS:ICD10CM:I25.11); or
 Atherosclerosis of native arteries of the extremities (UMLS:ICD10CM:I70.2); or
 Atherosclerosis of native arteries of extremities with intermittent claudication (UMLS:ICD10CM:I70.21); or
 Atherosclerosis of other arteries (UMLS:ICD10CM:I70.8); or
 Atherosclerosis of coronary artery bypass graft(s) without angina pectoris (UMLS:ICD10CM:I25.810); or
 Atherosclerosis of other coronary vessels without angina pectoris (UMLS:ICD10CM:I25.81); or
 Atherosclerotic heart disease of native coronary artery with unstable angina pectoris (UMLS:ICD10CM:I25.110).

Essential Hypertension
 Patients must have:
 Essential (primary) hypertension (UMLS:ICD10CM:I10).

Type 2 Diabetes Mellitus
 Patients must have:
 Type 2 diabetes mellitus (UMLS:ICD10CM:E11).

Aortic Aneurysm and Dissection
 Patients must have:
 Aortic aneurysm and dissection (UMLS:ICD10CM:I71).

Non-Traumatic Hemorrhagic Stroke
 Patients must have:
 Nontraumatic intracerebral hemorrhage (UMLS:ICD10CM:I61).

Peripheral Artery Disease
 Patients must have:
 any of the following:
 Other peripheral vascular diseases (UMLS:ICD10CM:I73); or
 Other peripheral vascular diseases (UMLS:ICD10CM:I73); or
 Atherosclerosis of native arteries of extremities with intermittent claudication, bilateral legs (UMLS:ICD10CM:I70.213); or
 Atherosclerosis of native arteries of extremities with intermittent claudication, other extremity (UMLS:ICD10CM:I70.218); or
 Atherosclerosis of native arteries of extremities with intermittent claudication, left leg (UMLS:ICD10CM:I70.212); or
 Atherosclerosis of native arteries of extremities with intermittent claudication, right leg (UMLS:ICD10CM:I70.211); or
 Atherosclerosis of native arteries of extremities with intermittent claudication, unspecified extremity (UMLS:ICD10CM:I70.219); or
 Atherosclerosis of native arteries of extremities with intermittent claudication (UMLS:ICD10CM:I70.21).

Gestational Diabetes
 Patients must have:
 any of the following:
 Gestational diabetes mellitus (UMLS:ICD10CM:O24.4); or
 Type 2 diabetes mellitus (UMLS:ICD10CM:E11); or
 Abnormal glucose complicating pregnancy (UMLS:ICD10CM:O99.810); or
 Unspecified diabetes mellitus in pregnancy, first trimester (UMLS:ICD10CM:O24.911); or
 Unspecified diabetes mellitus in pregnancy, unspecified trimester (UMLS:ICD10CM:O24.919); or
 Gestational diabetes mellitus in pregnancy, unspecified control (UMLS:ICD10CM:O24.419); or
 Gestational diabetes mellitus in childbirth, unspecified control (UMLS:ICD10CM:O24.429); or
 Gestational diabetes mellitus in the puerperium, unspecified control (UMLS:ICD10CM:O24.439); or
 Gestational diabetes mellitus in pregnancy (UMLS:ICD10CM:O24.41); or
 Gestational diabetes mellitus in childbirth (UMLS:ICD10CM:O24.42); or
 Gestational diabetes mellitus in the puerperium (UMLS:ICD10CM:O24.43); or
 Gestational diabetes mellitus in pregnancy, diet controlled (UMLS:ICD10CM:O24.410); or
 Gestational diabetes mellitus in pregnancy, insulin controlled (UMLS:ICD10CM:O24.414); or
 Gestational diabetes mellitus in childbirth, diet controlled (UMLS:ICD10CM:O24.420); or
 Gestational diabetes mellitus in childbirth, insulin controlled (UMLS:ICD10CM:O24.424); or
 Gestational diabetes mellitus in the puerperium, diet controlled (UMLS:ICD10CM:O24.430); or
 Gestational diabetes mellitus in the puerperium, insulin controlled (UMLS:ICD10CM:O24.434); or
 Abnormal glucose complicating pregnancy, childbirth and the puerperium (UMLS:ICD10CM:O99.81).

Insulin Resistance
 Patients must have:
 any of the following:
 Insulin resistance, unspecified (UMLS:ICD10CM:E88.819); or
 Other insulin resistance (UMLS:ICD10CM:E88.818); or
 Insulin resistance syndrome, Type A (UMLS:ICD10CM:E88.811); or
 Trimethylaminuria (UMLS:ICD10CM:E72.52).

Metabolic Syndrome
 Patients must have:
 any of the following:
 Metabolic syndrome (UMLS:ICD10CM:E88.810); or
 Metabolic syndrome and other insulin resistance (UMLS:ICD10CM:E88.81).

Dyslipidemia
 Patients must have:
 any of the following:
 Hyperlipidemia, unspecified (UMLS:ICD10CM:E78.5); or
 Other hyperlipidemia (UMLS:ICD10CM:E78.4); or
 Hyperlipidemia, unspecified (UMLS:ICD10CM:E78.5); or
 Other hyperlipidemia (UMLS:ICD10CM:E78.4); or
 Pure hypercholesterolemia (UMLS:ICD10CM:E78.0); or
 Mixed hyperlipidemia (UMLS:ICD10CM:E78.2); or
 Pure hyperglyceridemia (UMLS:ICD10CM:E78.1); or
 Hyperchylomicronemia (UMLS:ICD10CM:E78.3); or
 Other hyperlipidemia (UMLS:ICD10CM:E78.49); or
 Disorders of lipoprotein metabolism and other lipidemias (UMLS:ICD10CM:E78).

NAFLD
 Patients must have:
 any of the following:
 Fatty (change of) liver, not elsewhere classified (UMLS:ICD10CM:K76.0); or
 Nonalcoholic steatohepatitis (NASH) (UMLS:ICD10CM:K75.81); or
 Fibrosis and cirrhosis of liver (UMLS:ICD10CM:K74); or
 Hepatic fibrosis (UMLS:ICD10CM:K74.0).

CKD
 Patients must have:
 any of the following:
 Chronic kidney disease (CKD) (UMLS:ICD10CM:N18); or
 End stage renal disease (UMLS:ICD10CM:N18.6); or
 Chronic kidney disease, unspecified (UMLS:ICD10CM:N18.9); or
 Chronic kidney disease, stage 3 (moderate) (UMLS:ICD10CM:N18.3); or
 Chronic kidney disease, stage 4 (severe) (UMLS:ICD10CM:N18.4); or
 Chronic kidney disease, stage 5 (UMLS:ICD10CM:N18.5); or
 Chronic kidney disease, stage 1 (UMLS:ICD10CM:N18.1).

PCOS
 Patients must have:
 Polycystic ovarian syndrome (UMLS:ICD10CM:E28.2).

Systemic Autoimmune Connective Tissue Disorders
 Patients must have:
 Systemic connective tissue disorders (UMLS:ICD10CM:M30-M36).
